# Build fair machine learning models to predict adverse outcomes for Heart failure patients with preserved ejection fraction (HFpEF) and with reduced ejection fraction (HFrEF)

**DOI:** 10.64898/2025.12.18.25342417

**Authors:** YaYun Yeh, YaoAn Lee, Yu Huang, Wenxi Huang, Stephen E. Kimmel, Carl Yang, Zhe Jiang, Tingsong Xiao, Jiang Bian, Jingchuan Guo

**Affiliations:** Department of Pharmacy Practice, Purdue University College of Pharmacy, Indianapolis, Indiana, USA; Regenstrief Institute. Indianapolis, Indiana, USA; Biostatistics and Health Data Science. Indiana University, Indianapolis, Indiana, USA; Pharmaceutical Outcomes & Policy. University of Florida, Gainesville, Florida, USA; Department of Epidemiology. University of Florida, Gainesville, Florida, USA; Data and Information Systems Research Lab. Emory University, Atlanta, Georgia, USA; Department of Computer & Information Science & Engineering. University of Florida, Gainesville, Florida, USA

**Keywords:** heart failure with preserved ejection fraction (HFpEF), heart failure with reduced ejection fraction (HFrEF), Machine Learning (ML), Prediction, Fairness, Social Determinants of Health (SDoH)

## Abstract

**Background:** Heart failure (HF), including heart failure with preserved ejection fraction (HFpEF) and heart failure with reduced ejection fraction (HFrEF), remains a major global health challenge, particularly among aging populations. Timely and accurate prediction of severe adverse outcomes associated with HF is critical for optimizing care, reducing disease burden, and improving outcomes. Although social determinants of health (SDoH) have been recognized as key drivers of HF disparities and associated adverse outcomes, they are rarely integrated into HF prediction models, and fairness in such models remains understudied.

**Objective:** To develop and validate fairness-aware machine learning (ML) models incorporating both clinical and SDoH features to predict 6-month readmission or mortality in patients with HFpEF and HFrEF.

**Methods:** We conducted a retrospective cohort study using data from the University of Florida (UF) Health electronic health records (EHR). We included adult patients hospitalized for HF from 2016-2022 and followed up for 6 months to identify the incidence of readmission or mortality (a composite outcome). We developed machine learning models using logistic regression (LRC) and XGBoost algorithms, incorporating patients’ clinical characteristics, contextual SDoH (e.g., neighborhood deprivation index), and individual-level SDoH (extracted from clinical notes via natural language processing). Models were trained on balanced datasets using random oversampling, and performance was assessed via **C statistic**, F1-score, and recall. Fairness was evaluated using false negative rate (FNR) parity across sex, race/ethnicity, and age band. Bias mitigation strategies included Disparate Impact Remover, Adversarial Debiasing, and Calibrated Equalized Odds.

**Results:** HFpEF and HFrEF model that including both clinical and SDoH, achieved C statistic of 0.603 and 0.641, respectively in LRC, while clinical characteristics-only models had lower prediction performance (0.586 and 0.637, respectively). SHAP analysis identified strong predictors of sodium, financial constraint level and emergency department visit count for HFpEF cohort; inpatient visit count, financial constraint level and outpatient visit for HFrEF patients. Fairness assessment showed bias towards in HFpEF population, bias towards Black vs. White (FNR_black_ / FNR_white_= 0.7834) was increased by Disparate Impact Remover (0.8728). In HFrEF, bias towards Hispanic vs. White (FNR_hispanic_ / FNR_white_= 1.2217) was mitigated by Adversarial Debiasing (0.9880).

**Conclusion:** The prediction models for HFpEF and HFrEF offer an explainable and equity-enhancing approach for risk stratification in personalized HF clinical care. By integrating SDoH, our models’ show improved prediction utility and support targeted interventions for both medical and non-medical needs that are essential for patients’ health outcomes and critical for clinical decision-making.

## Introduction

Heart failure (HF) is a leading cause of hospitalization and mortality worldwide, which is classified into heart failure with preserved ejection fraction (HFpEF) and heart failure with reduced ejection fraction (HFrEF), each characterized by distinct pathophysiology, clinical features, and treatment approaches. HF affects more than 64 million individuals worldwide¹. In the United States, approximately 33.4% of HF patients have HFpEF, while 66.6% have HFrEF^2^. Worsening HF, marked by symptomatic and clinical decline, is observed in nearly one in six HFrEF patients within 18 months of diagnosis^3^. For HFpEF, limited treatment options and heterogeneous pathophysiology complicate clinical decision-making and risk stratification.

Therefore, accurate prediction of adverse outcomes in HF is essential for guiding treatment strategies, reducing hospital readmissions, and improving quality of life. Previous studies have identified several key predictive factors for HF-related adverse events. For example, elevated NT-proBNP levels, prior HF hospitalizations, diabetes mellitus, and older age were found to be strong independent predictors of all-cause mortality and hospitalization in patients with HFpEF^4^. In patients with HFrEF, risk factors for adverse outcomes include advanced age, renal dysfunction, anemia, and a history of myocardial infarction^5^. Other studies have identified demographic and clinical characteristics—including older age, female sex, and polypharmacy—as relevant predictors. A post hoc analysis of the TOPCAT trial demonstrated that hyper-polypharmacy was associated with an increased risk of hospitalization and serious adverse events in patients with HFpEF⁶. In addition, recent evidence points to contextual-level social determinants of health (SDoH), including housing instability, healthcare access, and environmental exposures, as contributing factors. Housing instability has been associated with increased emergency department use and delayed access to medical care, factors which may adversely affect chronic disease management such as in heart failure^7^. Healthcare utilization metrics such as frequent emergency department visits and hospital admissions have also been associated with worse outcomes. Patients with frequent Emergency department (ED) visits for acute HF accounted for over 55% of all HF-related hospitalizations, indicating a higher risk of adverse events⁸.

Significant disparities have been documented in both HFpEF and HFrEF and their clinical outcomes. For instance, Black patients are nearly 2.5 times more likely to be hospitalized for HF compared to White patients^9^. These disparities underscore the importance of addressing SDoH factors, including both contextual and individual SDoH, in understanding and managing HF. Contextual-level SDoH refer to factors measured from an individual’s surroundings, encompassing both social and physical environments, such as neighborhood characteristics, healthcare quality, and the built environment. Individual-level SDoH refer to personal social and economic conditions that influence health, such as income, education, employment status, housing stability, and access to transportation. Studies have shown that both contextual and individual-level SDoH contributed to disparities in HFpEF and HFrEF patient outcomes^9^.

Despite recognizing these predictors, integrating machine learning (ML)-based prediction models into clinical HF care remains challenging due to issues such as data overload, limited clinician capacity, and underutilization of social risk information. Few studies have incorporated both clinical and contextual-level SDoH in HF risk prediction models, which is particularly concerning given HF’s high burden, heterogeneity, and known disparities in outcomes across racial-ethnic and socioeconomic groups. A personalized and fair approach for risk management is urgently needed.

In this study, leveraging real world data (RWD) from the University of Florida (UF) Health’s EHRs, we incorporated both clinical and SDoH information and developed ML algorithms to predict adverse outcomes of HFpEF and HFrEF, respectively. We focused on the enhancement of model fairness and explainability to identify causal and modifiable risk factors that can be targeted for intervention.

## Methods

### Study Design and Population

We conducted a retrospective cohort study using UF Health Integrated Data Repository EHRs from 2016 to 2022. The cohort included adult patients diagnosed with HFpEF or HFrEF. Identification of HFpEF and HFrEF was based on validated algorithms with international classification of diseases (ICD) diagnosis codes^10^ (**Table S1**). We defined the index date as the first recorded inpatient encounter for HF between 2016 and 2022 and followed up patients for subsequent readmission or mortality. The workflow for cohort identification and machine learning model development is shown in **Figure S1**.

### Study outcome

The study outcome was the composite of readmission for HF or death within six months following the index date. Readmission was identified by subsequent inpatient encounter with a primary diagnosis of HF. Mortality was identified using the date of death recorded in the UF Health EHR, based on structured death date fields and discharge disposition status. Patients were censored at six months post-index or at the time of death, whichever occurred first.

### Predictor candidates

We collected a comprehensive set of demographics, clinical, and SDoH (contextual- and individual-level) variables at baseline to develop ML model for predicting 6-month HF readmission and death in HFpEF and HFrEF, respectively (**Table** S2 and **Table** S3). The variables were selected based on clinical expertise and prior literature^11^. Clinical variables were identified from structured EHR data using ICD diagnosis codes for comorbidities and RxNorm codes for medications. Individual-level SDoH variables were extracted from unstructured clinical notes using natural language processing (NLP)^12^, while contextual-level SDoH were derived from geospatially linked community-level datasets. All candidate predictor variables were collected from the pre-index baseline period and the index hospitalization, ensuring that only information available up to discharge was used as predictors.

### Development of ML Pipeline for HFpEF and HFrEF Prediction Models

The ML development process followed five main steps: data preprocessing, model training, performance evaluation, explanation, and fairness assessment. Two supervised ML algorithms, Logistic regression (LRC) and XGBoost, were developed independently for two datasets: HFpEF and HFrEF.

#### Step 1. Preprocessing

Missing values in continuous variables were imputed using median imputation, and mode imputation was applied for categorical variables. Features with high missingness or strong collinearity were excluded to improve robustness and reduce overfitting.

#### Step 2. ML modeling

The dataset was split randomly into training (70%), validation (10%), and testing (20%) sets. Random oversampling was used to address class imbalance within the training set. We trained two ML models, XGBoost and LRC, using grid search cross-validation to optimize hyperparameters.

#### Step 3. Performance Assessment

Model performance was assessed using multiple metrics, including the C statistic, F1-score, precision, recall, and specificity.

#### Step 4. Model Explanation

SHapley Additive exPlanations (SHAP) were applied for model interpretability. SHAP values helped rank the importance of both clinical and SDoH features in driving adverse outcomes in different HF subtypes. To further explore the complex interactions among features, we generated a causal graph using Mixed Graphical Models with the Conservative PC (CPC) algorithm, to identify important causal factors contributing to the outcome. This graph highlights the relationships between SDoH, comorbidities and clinical variables, with red edges indicating indirect pathways connecting SDoH to the outcome.

#### Step 5. Fairness Assessment and Bias Mitigation

In evaluating algorithmic fairness, we used the equality of opportunity metric, measured by the FNR (False negative rate) to assess fairness across subgroups defined by sex, race and ethnicity. The FNR represents the probability of incorrectly predicting no adverse event in patients who experienced one. To mitigate potential biases, we implemented three fairness-enhancing methods: pre-processing (Disparate Impact Remover), in-processing (Adversarial Debiasing), and post-processing (Calibrated Equalized Odds). Disparate Impact Remover weakens associations between features and protected attributes while preserving ranks. Adversarial Debiasing uses an adversary to discourage group dependent predictions and improve equalized odds. Calibrated Equalized Odds applied group specific thresholds to reduce true positive and false positive gaps while maintaining calibration. We report C statistic and FNR metrics before and after each mitigation method.

## Results

### Descriptive statistics of the study cohort

Our final cohort comprised 6,203 eligible patients hospitalized for HFpEF or HFrEF. **Table 1** shows the study cohort’s demographics and clinical characteristics.

**Table 1.** Demographic and Clinical Characteristics of the Study Cohort Stratified by 6-month readmission/death risk deciles (a.) HFpEF cohort (b.) HFrEF cohort.

| Name | top 10 decile | 10 - 50 decile | bottom 50 decile | p-value |
| --- | --- | --- | --- | --- |
| Age, years | 66.1 (13.1) | 65.6 (13.7) | 61.0 (14.0) | 0.816 |
| BMI (kg/m<sup>2</sup>) | 27.4 (7.2) | 29.6 (8.3) | 33.8 (10.9) | 0.01 |
| Weight (kg) | 81.8 (20.0) | 86.6 (24.6) | 98.9 (29.9) | 0.045 |
| Heart rate (beats/min) | 87.4 (16.3) | 81.9 (16.4) | 80.2 (15.9) | 0.683 |
| Systolic blood pressure (mmHg) | 116.6 (24.7) | 129.8 (23.8) | 141.9 (22.9) | <0.001 |
| Diastolic blood pressure (mmHg) | 67.4 (13.6) | 74.7 (14.7) | 82.4 (15.5) | 0.002 |
| Nitrate use | 45 (31.47%) | 110 (19.13%) | 68 (9.46%) | 0.01 |
| Positive inotropic agents | 9 (6.29%) | 23 (4.00%) | 5 (0.70%) | 1 |
| Loop diuretics | 79 (55.24%) | 179 (31.13%) | 97 (13.49%) | 0.002 |
| Non-loop diuretics | 52 (36.36%) | 73 (12.70%) | 44 (6.12%) | 0.935 |
| SGLT2 inhibitors | 0 (0.00%) | 4 (0.70%) | 0 (0.00%) | 1 |
| Beta-blockers | 93 (65.03%) | 207 (36.00%) | 147 (20.45%) | 0.022 |
| Calcium channel blockers | 40 (27.97%) | 118 (20.52%) | 89 (12.38%) | 0.988 |
| Non-insulin glucose-lowering drugs | 16 (11.19%) | 55 (9.57%) | 47 (6.54%) | 0.419 |
| Insulin use | 55 (38.46%) | 122 (21.22%) | 77 (10.71%) | 0.076 |
| Anticoagulants | 35 (24.48%) | 81 (14.09%) | 59 (8.21%) | 0.812 |
| Atrial fibrillation | 62 (43.36%) | 167 (29.04%) | 118 (16.41%) | 0.444 |
| Chronic obstructive pulmonary disease | 71 (49.65%) | 246 (42.78%) | 211 (29.35%) | 0.152 |
| Diabetes mellitus | 71 (49.65%) | 246 (42.78%) | 211 (29.35%) | 0.152 |
| Hypertension | 122 (85.31%) | 413 (71.83%) | 414 (57.58%) | 0.149 |
| Ischemic cardiovascular disease | 119 (83.22%) | 331 (57.57%) | 233 (32.41%) | 0.027 |
| History of myocardial infarction | 49 (34.27%) | 137 (23.83%) | 66 (9.18%) | 0.608 |
| Chronic kidney disease | 0 (0.00%) | 0 (0.00%) | 0 (0.00%) | 1 |
| Anemia | 82 (57.34%) | 235 (40.87%) | 157 (21.84%) | 0.006 |
| Obesity | 27 (18.88%) | 142 (24.70%) | 175 (24.34%) | 1 |
| Serum creatinine (mg/dL) | 1.8 (1.1) | 2.0 (2.1) | 1.3 (1.4) | 0.453 |
| LDL cholesterol (mg/dL) | 73.2 (35.8) | 84.7 (37.0) | 92.0 (40.6) | 0.742 |
| HDL cholesterol (mg/dL) | 43.3 (16.8) | 46.2 (16.6) | 48.7 (19.1) | 0.52 |
| Hemoglobin (g/dL) | 6.5 (1.7) | 6.7 (1.8) | 7.1 (2.3) | 0.536 |

\*All comorbidities and medication variables are binary indicators (1 = yes, 0 = no).

(b.) HFrEF cohort
| Name | top 10 decile | 10 - 50 decile | bottom 50 decile | p-value |
| --- | --- | --- | --- | --- |
| Name | 66.1 (13.1) | 65.6 (13.7) | 61.0 (14.0) | 0.816 |
| Age, years | 27.4 (7.2) | 29.6 (8.3) | 33.8 (10.9) | 0.01 |
| BMI (kg/m<sup>2</sup>) | 81.8 (20.0) | 86.6 (24.6) | 98.9 (29.9) | 0.045 |
| Weight (kg) | 87.4 (16.3) | 81.9 (16.4) | 80.2 (15.9) | 0.683 |
| Heart rate (beats/min) | 116.6 (24.7) | 129.8 (23.8) | 141.9 (22.9) | <0.001 |
| Systolic blood pressure (mmHg) | 67.4 (13.6) | 74.7 (14.7) | 82.4 (15.5) | 0.002 |
| Diastolic blood pressure (mmHg) | 45 (31.47%) | 110 (19.13%) | 68 (9.46%) | 0.01 |
| Nitrate use | 9 (6.29%) | 23 (4.00%) | 5 (0.70%) | 1 |
| Positive inotropic agents | 79 (55.24%) | 179 (31.13%) | 97 (13.49%) | 0.002 |
| Loop diuretics | 52 (36.36%) | 73 (12.70%) | 44 (6.12%) | 0.935 |
| Non-loop diuretics | 0 (0.00%) | 4 (0.70%) | 0 (0.00%) | 1 |
| SGLT2 inhibitors | 93 (65.03%) | 207 (36.00%) | 147 (20.45%) | 0.022 |
| Beta-blockers | 40 (27.97%) | 118 (20.52%) | 89 (12.38%) | 0.988 |
| Calcium channel blockers | 16 (11.19%) | 55 (9.57%) | 47 (6.54%) | 0.419 |
| Non-insulin glucose-lowering drugs | 55 (38.46%) | 122 (21.22%) | 77 (10.71%) | 0.076 |
| Insulin use | 35 (24.48%) | 81 (14.09%) | 59 (8.21%) | 0.812 |
| Anticoagulants | 62 (43.36%) | 167 (29.04%) | 118 (16.41%) | 0.444 |
| Atrial fibrillation | 71 (49.65%) | 246 (42.78%) | 211 (29.35%) | 0.152 |
| Chronic obstructive pulmonary disease | 71 (49.65%) | 246 (42.78%) | 211 (29.35%) | 0.152 |
| Diabetes mellitus | 122 (85.31%) | 413 (71.83%) | 414 (57.58%) | 0.149 |
| Hypertension | 119 (83.22%) | 331 (57.57%) | 233 (32.41%) | 0.027 |
| Ischemic cardiovascular disease | 49 (34.27%) | 137 (23.83%) | 66 (9.18%) | 0.608 |
| History of myocardial infarction | 0 (0.00%) | 0 (0.00%) | 0 (0.00%) | 1 |
| Chronic kidney disease | 82 (57.34%) | 235 (40.87%) | 157 (21.84%) | 0.006 |
| Anemia | 27 (18.88%) | 142 (24.70%) | 175 (24.34%) | 1 |
| Obesity | 1.8 (1.1) | 2.0 (2.1) | 1.3 (1.4) | 0.453 |
| Serum creatinine (mg/dL) | 73.2 (35.8) | 84.7 (37.0) | 92.0 (40.6) | 0.742 |
| LDL cholesterol (mg/dL) | 43.3 (16.8) | 46.2 (16.6) | 48.7 (19.1) | 0.52 |
| HDL cholesterol (mg/dL) | 6.5 (1.7) | 6.7 (1.8) | 7.1 (2.3) | 0.536 |
| Hemoglobin (g/dL) | 65.1 (277.8) | 129.9 (423.4) | 272.7 (1238.0) | 0.236 |

\*All comorbidities and medication variables are binary indicators (1 = yes, 0 = no).

Of the cohort, the mean age was 64.0 ± 14.1 years, 43.4% were females, 53.26% were NHW, 40.12% were NHB, 26.36% were Hispanic, 3.17% were other races and ethnicities, and 3.04% were unknown. NHB and Hispanic patients were generally younger than NHW patients (60.81 and 61.92 years vs. 66.48 years, respectively). Among the HFpEF cohort, hypertension (79.3%), COPD (54.2%), and diabetes (54.2%) were most prevalent; in the HFrEF cohort, hypertension (67.3%), ischemic cardiovascular disease (46.6%), and COPD (38.6%) were most prevalent.

### ML prediction models in HFpEF and HFrEF

We compared three sets of model performance: clinical only, clinical with individual SDoH and full variables (clinical plus both individual and contextual SDoH.) For LRC model in HFpEF cohort, the C statistic were 0.5863, 0.5805, and 0.6032 (**Table** S4; in HFrEF were 0.637, 0.6486 and 0.6407. Adding individual SDoH to clinical variables yielded the largest numerical gain in HFrEF cohort, whereas adding contextual SDoH on top of clinical plus individual SDoH yielded the largest gain in HFpEF cohort. Because LRC achieved comparable discrimination to XGBoost and offers more stable and interpretable coefficients, we used the LRC models as the primary models for SHAP-based explainability and causal discovery analyses. **Figure 1** presents the receiver operating characteristic (ROC) curves illustrating the performance of LRC and XGBoost prediction models with random oversampling.

**Figure 1.**
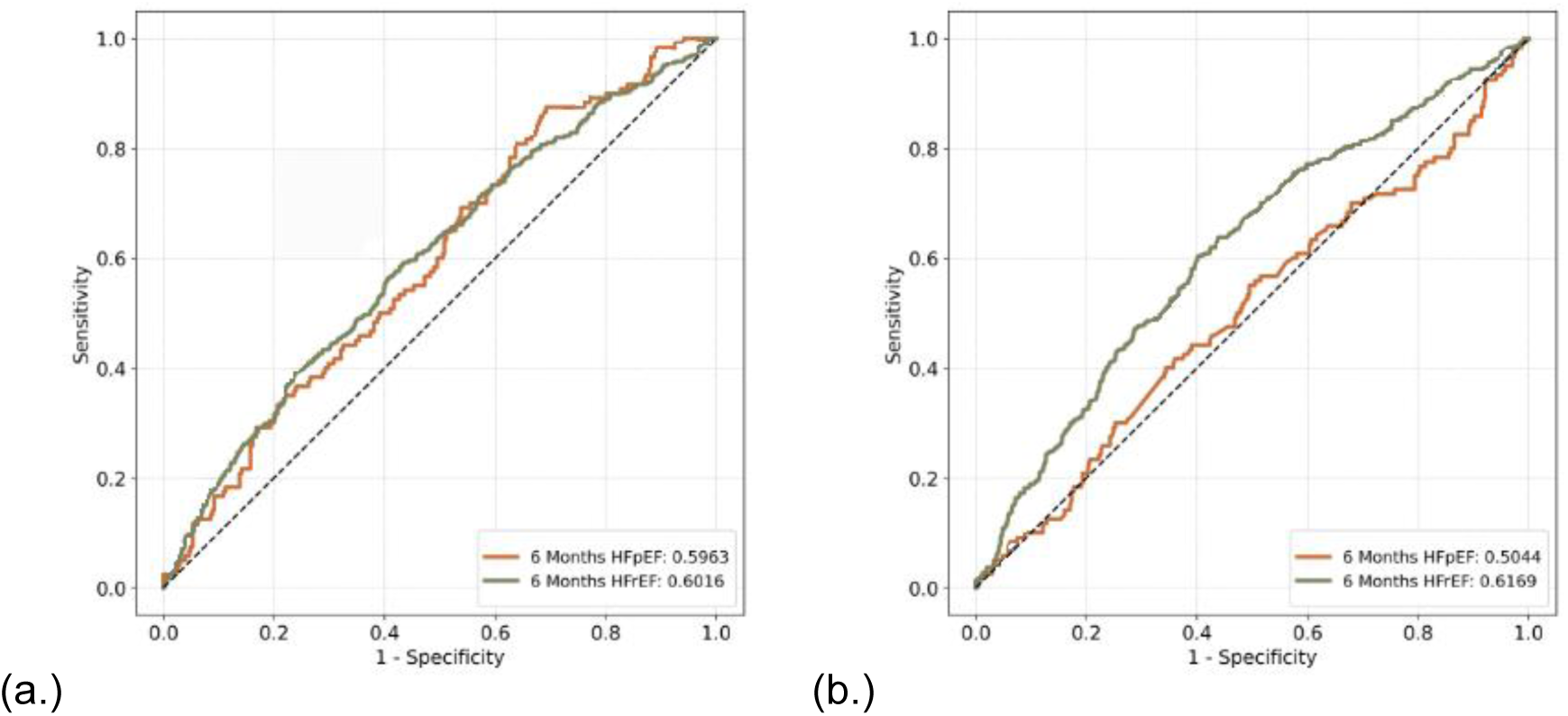
Model performance of LRC. Each plot shows ROC curves for models trained on HFpEF-cohort, HFrEF-cohort

In the independent evaluation dataset, we stratified the predicted 6-month risk of readmission or death by decile using the ML generated risk score, demonstrating excellent calibration utility in identifying HF individuals at risk of adverse outcomes. For example, in the LRC HFpEF group, the 6-month risk of HF adverse outcomes in the top 20% of patient cohort was 40%, more than two times that of the bottom decile (**Figure** 2a). Similarly, in XGBoost model (Figure S2), the top 20% predicted cased accounted for 41.46% of the 6-month adverse outcome risk.

**Figure 2.**
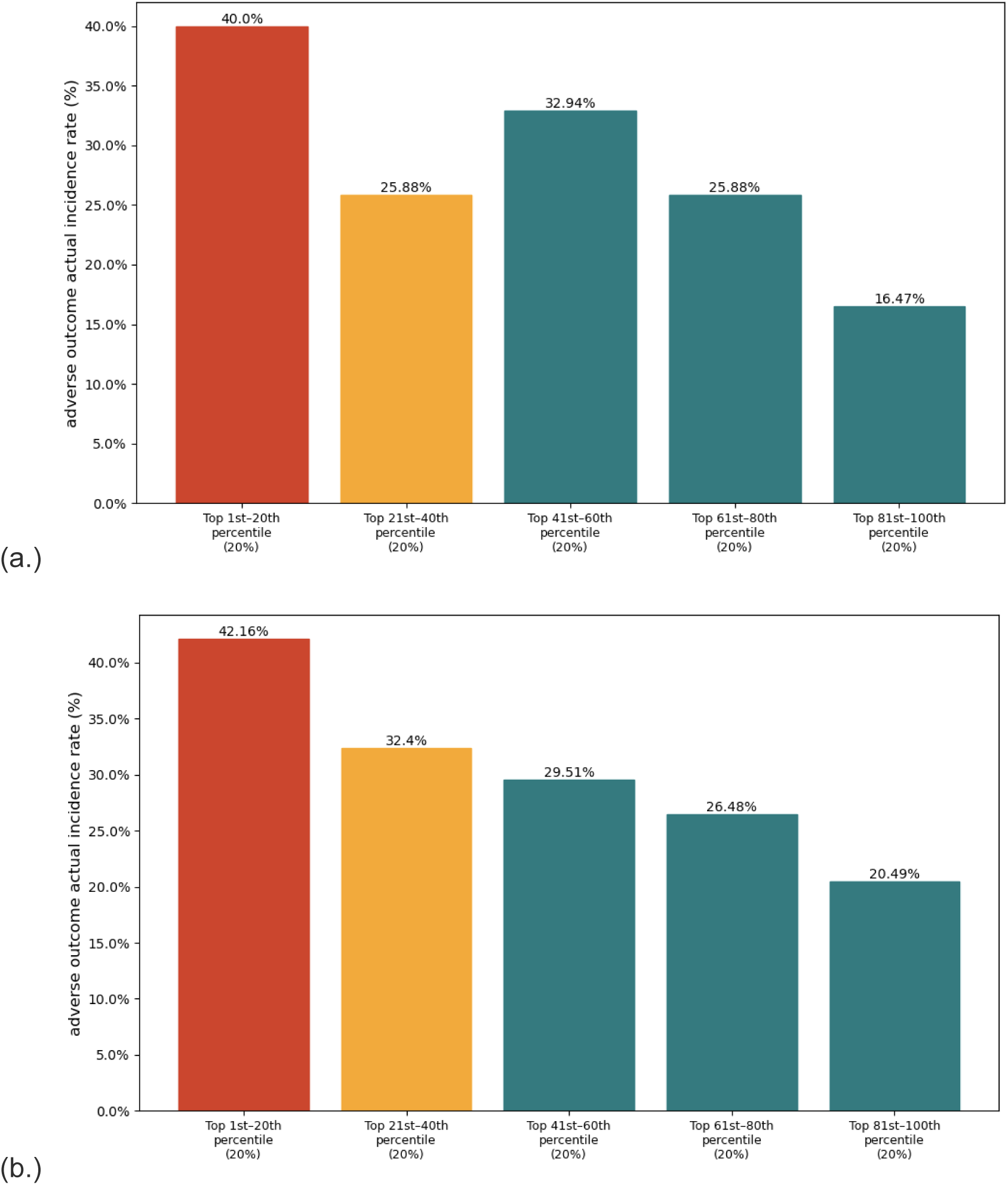
6-month readmission/death risk by machine learning prediction risk decile using LRC model. (a.) HFpEF cohort (b.) HFrEF cohort

### Explainable AI to identify important features contributing to ML models predicting adverse outcomes in HFpEF and HFrEF

The two HF subtypes identified different sets of important features for predicting HF readmission and mortality outcomes in LRC model (**Figure** 3). In HFpEF, sodium, financial constraint level and emergency department visit count were the top three most important predictors, while in the HFrEF cohort, inpatient visit count, financial constraint level and outpatient visit count were identified as most important predictions. In contrast, the XGBoost model (**Figure** S3) assigned a different relative importance to these variables’ serum albumin, financial constraint level, Age at index data for HFpEF. inpatient visit count, systolic blood pressure, and financial constraint level, which was identified as important in model’s prediction of adverse outcomes.

**Figure 3.**
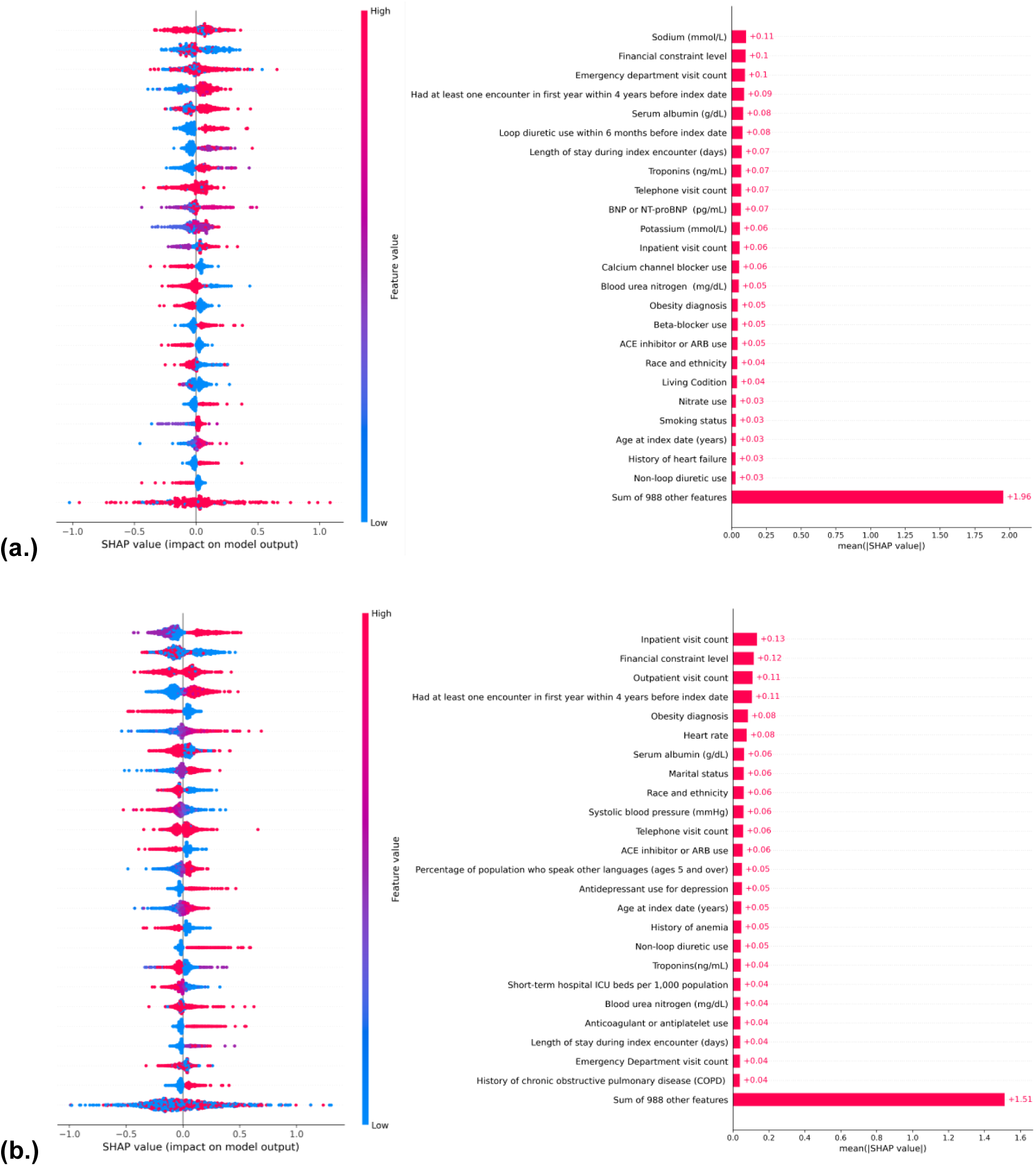
SHAP values of important predictions from the original LRC. (a.) HFpEF cohort (b.) HFrEF cohort.

**Figure** 4 Causal structure learning results using the CPC algorithm across two different HF cohorts: (a.) HFpEF and (b.) HFrEF. These two graphs represent our exploratory analysis using causal discovery to identify potential relationships among demographic, clinical and SDoH features. In LRC model HFpEF population cohort, we identified 1 demographic variable, 9 clinical variables, 7 comorbidities, 2 individual level SDoH variables. The resulting causal graph included 20 nodes (including the outcome) and 23 edges, representing direct or indirect relationships among these variables. In the HFrEF-only cohort, we identified 1 demographic variable, 6 clinical variables, 5 comorbidities, and 1 individual-level SDoH. The resulting causal graph included 16 nodes and 28 edges. Notably, two indirect relationships between SDoH variables and the outcome were observed in HFpEF cohorts, while one indirect relationship was identified in the HFrEF cohort. Among SDoH variables, financial constrain was found to be causally related to adverse outcome in patient with HFpEF and HFrEF, either directly or indirectly in both models LRC and XGBoost (**Figure S4**).

**Figure 4.**
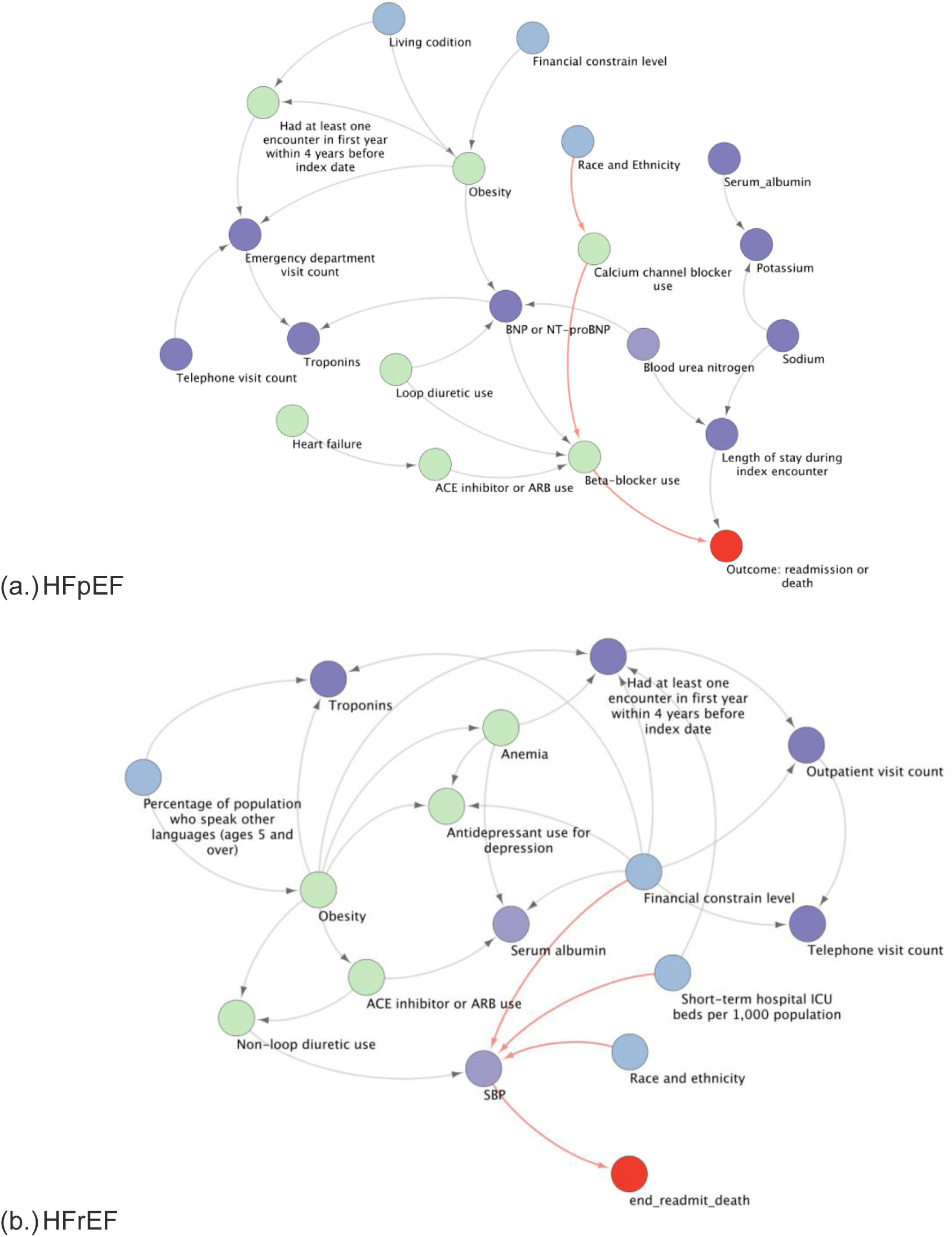
The causal discovery results on full data with xgboost model from SHAP analysis. These three images are results from CPC models. The blue nodes present SDoH and demographics variables, the green nodes stand for comorbidities and medication variables, purple nodes stand for the clinical variables, and the red node indicates the outcome. The red edges represent the indirect relationships between SDoH and outcome. (a.) HFpEF cohort (b.) HFrEF cohort

### Fairness assessment and mitigation

We assessed fairness in our ML models by evaluating FNR parity across subgroups by sex (male, female), race/ethnicity (Hispanic, White; Black, White). An FNR ratio greater than 1 indicates under-identification in the subgroup. A detailed summary of FNR ratios across subgroups and models is provided in **Table** S5. The corresponding subgroup FNR curves for these groups in the LRC and XGBoost models (**Figure** 5), where closer overlap reflects better parity.

**Figure 5.**
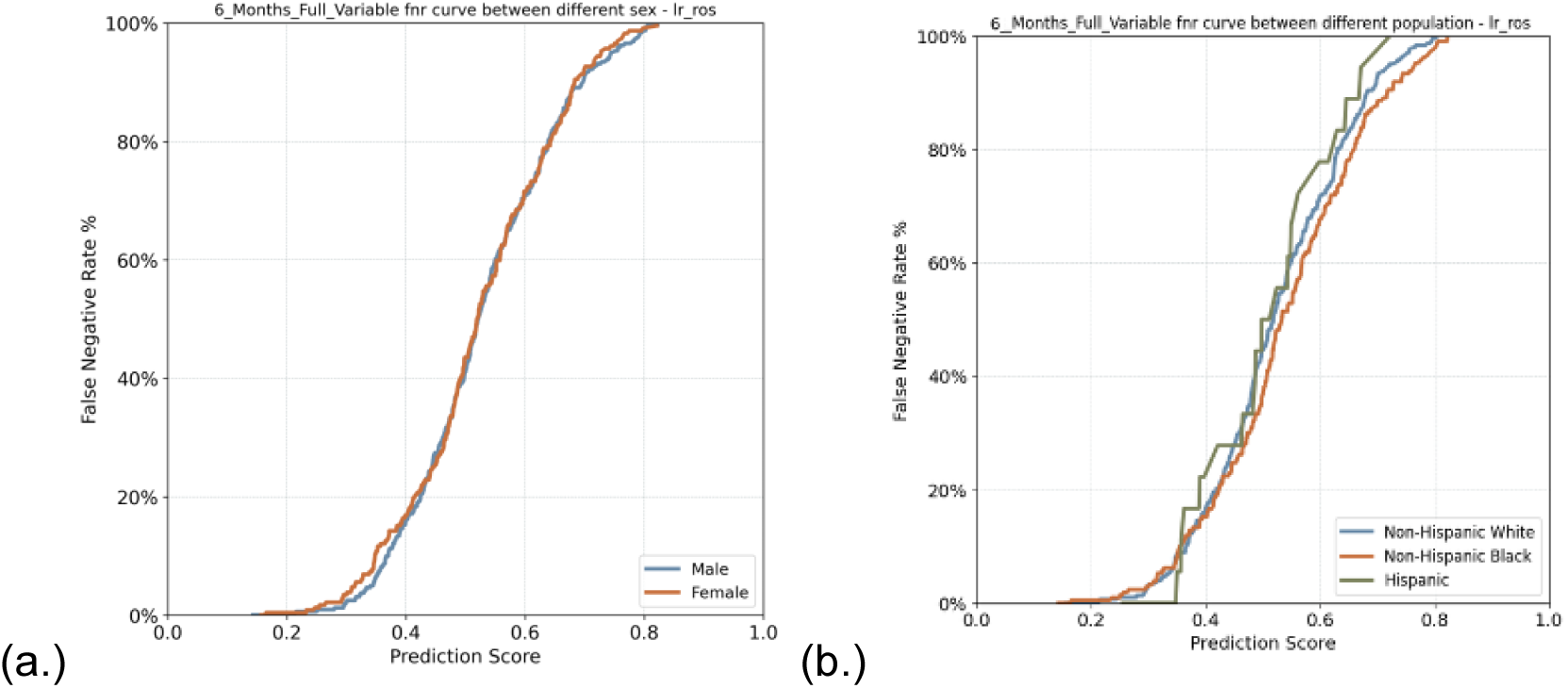
Evaluation of algorithm fairness with a focus on FNR disparity LRC (a.) by sex (b.) by non-Hispanic white, non-Hispanic black and Hispanic

A notable disparity was observed in HFpEF subtype, a bias towards Black patients (FNR_black_ / FNR_white_= 0.7834), indicating fewer missed Black than White patients, after applying Disparate Impact Remover, it was improved to (FNR_black_ / FNR_white_= 0.8728). In HFrEF subtype, the Hispanic-to-White FNR ratio was (FNR_hispanic_ / FNR_white_= 1.2217), indicating bias towards Hispanic patients, it was mitigated by Adversarial Debiasing (FNR_hispanic_ / FNR_white_= 0.9880), approximately parity.

To address these biases, we applied three fairness-enhancing techniques: Disparate Impact Remover, Adversarial Debiasing, and Calibrated Equalized Odds. All of which improved FNR parity, though with some trade-offs in model discrimination (C statistic) (**Figure** S6). Calibration curves also demonstrated improved alignment between predicted and observed risks across demographic subgroups, indicating greater reliability of the mitigated models.

## Discussion

In this study, we developed subtype-specific machine learning models to predict six-month adverse outcomes in patients with HFpEF and HFrEF, with careful attention to algorithmic fairness by applying fairness-aware mitigation techniques to enhance model explainability through SHAP values and causal inference via CPC algorithms. We found substantial differences in risk predictors and performance between the HF subtypes: the top 20% of patients identified by our models accounted for 40% of adverse events in HFpEF and 41.46% in HFrEF. Integrating clinical variables with individual- and contextual-level SDoH resulted in modest but meaningful improvements in model accuracy. Fairness assessments identified and effectively mitigated disparities in predictions related to sex and race/ethnicity through targeted fairness-aware methods. These results highlight the critical importance of tailored, explainable, and equitable prediction models that incorporate both clinical and social factors to improve clinical decision-making and health outcomes for diverse heart failure populations.

To our knowledge, this is among the first studies to develop HF subtype-specific prediction models of adverse outcomes among HFrEF and HFpEF patients, with careful consideration of algorithmic fairness and model explainability. We conducted separate models for HFpEF and HFrEF because these syndromes differ fundamentally in their underlying pathogenesis, risk factor profiles, disease trajectories, and therapeutic strategies. HFpEF is characterized by diastolic dysfunction, systemic inflammation, and metabolic comorbidities, whereas HFrEF involves systolic impairment, neurohormonal activation, and more established guideline directed medical therapies. A general approach would therefore obscure these subtype specific drivers of risk. The clinical implications of our models are substantial. The model effectively identified patients at highest risk for adverse outcomes. In the HFpEF cohort, the top 20% of patients by predicted risk accounted for 40% of six-month adverse events, whereas in the HFrEF cohort it captured 41.46%. Such subtype-specific stratification underscores the model’s ability to pinpoint those most likely to experience readmission or death and highlights the value of tailored interventions for both HFpEF and HFrEF patients (**Figure 2**).

Another unique aspect of the study is that we included not only clinical factors but also individual- and contextual- level SDoH for the prediction of adverse outcomes in both HFrEF and HFpEF. Health disparities in both HFrEF and HFpEF are significant, particularly among individuals from socially and economically disadvantaged populations. For example, studies have shown that social determinants such as food insecurity, housing instability, and limited access to transportation are independently associated with higher rates of hospitalization and mortality in HF patients, and that incorporating measures of neighborhood deprivation and individual-level socioeconomic status into risk models uncovers vulnerabilities not captured by clinical variables alone^14^. In our study, incorporating SDoH variables slightly improved model performance compared to clinical characteristics alone. Across all two cohorts, the full feature set (clinical, individual, and contextual SDoH) yielded small increases in the C statistic over clinical-only models (HFpEF 0.5863 to 0.6032, HFrEF 0.6370 to 0.6407. In HFrEF the gain was driven primarily by individual-level SDoH from 0.6370 to 0.6486. In HFpEF, C statistic increased from 0.5863 to 0.6032 when individual and contextual SDoH were added. Although these differences in C statistic are modest, SDoH contributed complementary information, highlighting non-clinical drivers of risk and supporting more equitable identification of high-risk patients in cohorts assessments. Contextual factors were particularly predictive in HFpEF patients, whereas individual-level SDoH contributed most of the gain in HFrEF patients. These findings imply the need for subtype-specific risk models that incorporate social and environmental context.

Our prediction models offer enhanced explainability, strengthening their clinical implications. By incorporating explainable AI tools such as SHAP values and causal structure learning techniques, CPC algorithms, our models provide interpretable insights into risk prediction.

Distinct key predictors were identified for HFrEF (e.g., BMI, left ventricular ejection fraction, diastolic blood pressure, and inpatient visit count) and HFpEF (e.g., length of stay at index encounter, BMI, diastolic blood pressure, serum albumin, and triglycerides). The causal structure learning framework additionally revealed potential causal links between SDoH (e.g., percentage of Medicare-only insured individuals, percentage of American Indian and Alaska native population, and neighborhood premature death rate) and clinical variables leading to adverse outcomes in HFrEF and HFpEF, suggesting the importance of integrating social risk management into the clinical care of HF (**Figure** 4 and **Figure** S4).

Our fairness assessment revealed robust performance across racial and ethnic groups, with FNR parity metrics within acceptable statistical fairness ranges (0.80 to 1.25). Nevertheless, we identified subtype-specific disparities between Black and White patients in HFpEF (FNR_Black_/FNR_White_ = 0.7834), and Hispanic and White patients in HFrEF (FNR_Hispanic_/FNR_White_ = 1.2217). These findings highlight that fairness challenges are context-dependent and require tailored mitigation strategies. We addressed these disparities using three fairness-aware ML techniques: Disparate Impact Remover, Adversarial Debiasing, and Calibrated Equalized Odds. Each method effectively reduced subtype-specific disparities, though slightly compromising C statistic performance, reflecting inherent trade-offs between fairness and accuracy. Further validation in larger, multi-institutional cohorts is needed to refine and generalize these fairness strategies.

Our study is subject to several limitations. Cohort identification via structured EHR diagnosis codes may include misclassified or false-positive cases, especially in non-specific HF presentations. Future studies should incorporate NLP-driven phenotype refinement for improved accuracy. Although UF Health provides diverse patient data, findings may not generalize broadly. Validation through federated learning and multi-center datasets is warranted. Our individual-level SDoH variables were limited to EHR-available data, potentially overlooking critical factors like income instability or caregiving stress; future work should integrate validated screening tools and community-level data. Mortality ascertainment relied on deaths recorded with in the UF Health EHR. Deaths occurring outside the system were likely missed, which could underestimate event rates and affect model calibration and discrimination. Additionally, potential overlaps between predictors and outcome definitions (e.g., prior hospitalization predicting readmission) may affect causal inference. Sensitivity analyses excluding overlapping predictors would strengthen future studies.

## Conclusion

The prediction models for HFpEF and HFrEF offer an explainable and equity-enhancing approach for risk stratification in personalized HF clinical care. By integrating SDoH into adverse outcome prediction modeling, our models support targeted interventions for both medical and non-medical needs that are essential for patients’ health outcomes and critical for clinical decision-making.

## Data Availability

The data used in this study are derived from UF Health electronic health records. Due to privacy regulations and institutional data use restrictions, the individual-level data are not publicly available.

## Notes

### Competing Interest Statement

The authors have declared no competing interest.

### Funding Statement

This research received no external funding.

### Author Declarations

Ethics committee/IRB of Full UF gave ethical approval for this work

