## Supplemental tables and figures for "Build fair machine learning models to predict adverse outcomes for Heart failure patients with preserved ejection fraction (HFpEF) and with reduced ejection fraction (HFrEF)"

**Supplemental materials**

**Table S1.** ICD Codes for HF Definitions

**Table S2.** Summary of Variables by Category

**Table S3.** Summary of Contextual-level SDoH

**Table S4.** Performance metrics for XGBoost and Logistic regression

**Table S5.** Opportunity of equality measured by false negative rate by different models on various feature sets

**Figure S1.** Workflow to identify cohort and build up ML model.

**Figure S2.** 6-month readmission/death risk by machine learning prediction risk decile using XGboost model. (a.) HFpEF population (b.) HFrEF population

**Figure S3**. SHAP values of important predictions from the original XGboost. (a.) HFpEF (b.) HFrEF.

**Figure S4.** The causal discovery results on full data with XGBoost model from SHAP analysis. These three images are results from CPC models. The blue nodes present SDoH and demographics variables, the green nodes stand for comorbidities and medication variables, purple nodes stand for the clinical variables, and the red node indicates the outcome. The red edges represent the indirect relationships between SDoH and outcome. (a.) HFpEF (b.) HFrEF

**Figure S5.** Impact of bias mitigation on Model Performance. The x-axis shows Equality of Opportunity, and the y-axis shows C statistic. Mitigation models reduced bias but also lowered C statistic (a.) FNR Ratio: Black/White in HFpEF (b.) FNR Ratio: Hispanic/White in HFrEF population.

**Figure S6.** Impact of bias mitigation on Model Performance. The x-axis shows Equality of Opportunity, and the y-axis shows C statistic. Mitigation models reduced bias but also lowered

C statistic (a.) FNR Ratio: Black/White in HFpEF (b.) FNR Ratio: Hispanic/White in HFrEF population.

**Table S1.** ICD Codes for Heart Failure

| Condition | | |
|---|---|---|
| HF | 402.x1 (402.01, 402.11, 402.91), 404.x1 (404.01, 404.11, 404.91), 404.x3 (404.03, 404.13, 404.93), 428, 428.x, 398.91 | I50, I50.x, I11.0, I13.0, I13.2, I97.13, I09.81 |
| HFpEF | 428.3x | I50.3x |
| HFrEF | 428.2x | I50.2x |

**Table S2.** Summary of Variables by Category

| Category | Variables list |
|---|---|
| Demographics | AGE, RACE_ETHNICITY, SEX |
| Clinical | Length of stay, Mechanical ventilation, HFpEF, heart failure history, Atrial fibrillation and/or flutter , COPD, Diabetes, Hypertension, Dyslipidemia, Ischemic cardiovascular disease, History of myocardial infarction, History of Stroke/TIA, Chronic kidney disease , Dialysis , Anemia, |

| | |
|---|---|
| | Depression , Cardiac and vascular device, Obesity , ICD and/or CRT-D Pacemaker and/or CRT-P, BMI, Weight, SBP, DBP, Heart rate, Female/Male Breast Cancer , Colorectal Cancer , Prostate Cancer , Lung Cancer, Endometrial Cancer, inpatient count, outpatient count, ED count |
| Medication | Sacubitril/Valsartan_baseline, SGLT2 inhibitors, Loop diuretics, non-loop diuretics, ACEIs, ARBs, Beta blockers, Calcium channel blockers, Positive inotropic agents, Nitrates, Statins, Insulin (use any type of insulin), non-insulin glucose lowering meds (any use: metformin, DPP-4, sulfonylureas, TZD, SGLT2i and GLP1a), Anticoagulants (warfarin and DOACs), Antiplatelets, Antidepressants |
| Laboratory | Hemoglobin, Lymphocyte count, BNP / NT-proBNP, Troponins/ High-sensitivity troponin, Total cholesterol, LDL, HDL, Triglycerides, eGFR, Serum creatinine, Blood urea nitrogen, Sodium, Potassium, Serum albumin |
| Individual SDoH | Education, occupation, Financial constrain, living condition, living supply, marital status, smoking, alcohol, drug use |

| | |
|---|---|
| Contextual SDoH | See supplementary table S3 |

**Table S3.** Summary of Contextual-level SDoH

| Category | Data source | Time period | Number of variables |
|---|---|---|---|
| Economic Stability | Agency for Healthcare Research and Quality | 2009-2019 | 88 |
| | County Health roadmap | 2009-2019 | 10 |
| | Local area unemployment statistics | 2019 | 4 |
| | Economic resilience | 2015 | 3 |
| Education Access and Quality | Agency for Healthcare Research and Quality | 2009-2019 | 19 |
| | County Health roadmap | 2009-2019 | 2 |
| Health Care Access and Quality | US Cancer statistics | 2018 | 8 |
| | Agency for Healthcare Research and Quality | 2009-2019 | 451 |
| | County Health roadmap | 2009-2019 | 54 |
| | Dartmouth health | 2009-2019 | 8 |
| Natural Environment | Agency for Healthcare Research and Quality | 2009-2019 | 97 |
| Neighborhood and Built Environment | Agency for Healthcare Research and Quality | 2009-2019 | 117 |

| | | | |
|---|---|---|---|
| | Air Quality Index | 2009-2019 | 15 |
| | Air Quality System | 2009-2019 | 14 |
| | County Health Roadmap | 2009-2019 | 18 |
| | Local Area Unemployment Statistics | 2009-2019 | 2 |
| | Religion | 2010 | 3 |
| | Water | 2017 | 242 |
| Social and Community Context | Social capital | 2009-2014 | 13 |
| | Agency for Healthcare Research and Quality | 2009-2019 | 138 |
| | County Health roadmap | 2009-2019 | 2 |

* All data sources are spatially scaled at the county level.

**Table S4.** Performance metrics for XGBoost and Logistic

| | | | F1-Score | C Statistic | Recall | Precision | Specificity |
|---|---|---|---|---|---|---|---|
| XGBoost | full variable | | | | | | |
| | HFpEF | mean | 0.370 | 0.550 | 0.463 | 0.321 | 0.604 |
| | | std.dev | 0.056 | 0.045 | 0.136 | 0.048 | 0.139 |
| | HFrEF | mean | 0.427 | 0.596 | 0.481 | 0.388 | 0.671 |
| | | std.dev | 0.029 | 0.024 | 0.061 | 0.026 | 0.058 |
| | Contextual SDoH | | | | | | |
| | HFpEF | mean | 0.380 | 0.528 | 0.506 | 0.307 | 0.550 |
| | | std.dev | 0.040 | 0.037 | 0.085 | 0.027 | 0.064 |
| | HFrEF | mean | 0.331 | 0.493 | 0.381 | 0.300 | 0.613 |
| | | std.dev | 0.03 | 0.024 | 0.086 | 0.023 | 0.094 |

| | | | | | | | |
|---|---|---|---|---|---|---|---|
| | Clinical | | | | | | |
| | HFpEF | mean | 0.364 | 0.551 | 0.433 | 0.322 | 0.635 |
| | | std.dev | 0.051 | 0.048 | 0.104 | 0.044 | 0.101 |
| | HFrEF | mean | 0.451 | 0.62 | 0.536 | 0.391 | 0.638 |
| | | std.dev | 0.030 | 0.030 | 0.055 | 0.027 | 0.048 |
| | Individual SDoH | | | | | | |
| | HFpEF | mean | 0.396 | 0.552 | 0.553 | 0.319 | 0.52 |
| | | std.dev | 0.052 | 0.045 | 0.162 | 0.036 | 0.171 |
| | HFrEF | mean | 0.414 | 0.562 | 0.53 | 0.341 | 0.556 |
| | | std.dev | 0.025 | 0.021 | 0.068 | 0.018 | 0.063 |
| | Individual SDoH + Contextual SDoH | | | | | | |
| | HFpEF | mean | 0.369 | 0.551 | 0.439 | 0.323 | 0.63 |
| | | std.dev | 0.048 | 0.036 | 0.088 | 0.034 | 0.063 |
| | HFrEF | mean | 0.375 | 0.524 | 0.458 | 0.323 | 0.584 |
| | | std.dev | 0.037 | 0.024 | 0.097 | 0.024 | 0.103 |
| | Clinical+ Individual SDoH | | | | | | |
| | HFpEF | mean | 0.330 | 0.563 | 0.334 | 0.340 | 0.743 |
| | | std.dev | 0.069 | 0.046 | 0.100 | 0.056 | 0.072 |
| | HFrEF | mean | 0.454 | 0.629 | 0.524 | 0.402 | 0.662 |
| | | std.dev | 0.031 | 0.026 | 0.057 | 0.028 | 0.047 |
| LRC | full variable | | | | | | |
| | HFpEF | mean | 0.416 | 0.603 | 0.494 | 0.360 | 0.654 |
| | | std.dev | 0.045 | 0.040 | 0.067 | 0.038 | 0.040 |
| | HFrEF | mean | 0.470 | 0.640 | 0.558 | 0.407 | 0.649 |
| | | std.dev | 0.021 | 0.019 | 0.033 | 0.018 | 0.022 |

| | | | | | | | |
|---|---|---|---|---|---|---|---|
| | Contextual SDoH | | | | | | |
| | HFpEF | mean | 0.382 | 0.541 | 0.501 | 0.310 | 0.561 |
| | | std.dev | 0.044 | 0.044 | 0.078 | 0.033 | 0.055 |
| | HFrEF | mean | 0.236 | 0.500 | 0.510 | 0.153 | 0.490 |
| | | std.dev | 0.232 | 0.000 | 0.502 | 0.151 | 0.502 |
| | Clinical | | | | | | |
| | HFpEF | mean | 0.396 | 0.586 | 0.475 | 0.342 | 0.640 |
| | | std.dev | 0.051 | 0.041 | 0.080 | 0.040 | 0.043 |
| | HFrEF | mean | 0.461 | 0.637 | 0.534 | 0.406 | 0.662 |
| | | std.dev | 0.021 | 0.018 | 0.032 | 0.019 | 0.023 |
| | Individual SDoH | | | | | | |
| | HFpEF | mean | 0.395 | 0.573 | 0.506 | 0.326 | 0.586 |
| | | std.dev | 0.045 | 0.044 | 0.072 | 0.036 | 0.049 |
| | HFrEF | mean | 0.432 | 0.585 | 0.550 | 0.357 | 0.572 |
| | | std.dev | 0.021 | 0.021 | 0.035 | 0.017 | 0.028 |
| | Individual SDoH+ Contextual SDoH | | | | | | |
| | HFpEF | mean | 0.393 | 0.559 | 0.510 | 0.322 | 0.575 |
| | | std.dev | 0.042 | 0.041 | 0.076 | 0.032 | 0.055 |
| | HFrEF | mean | 0.424 | 0.577 | 0.527 | 0.356 | 0.589 |
| | | std.dev | 0.023 | 0.021 | 0.042 | 0.018 | 0.033 |
| | Clinical+ Individual SDoH | | | | | | |
| | HFpEF | mean | 0.400 | 0.580 | 0.491 | 0.339 | 0.622 |
| | | std.dev | 0.047 | 0.043 | 0.078 | 0.037 | 0.049 |
| | HFrEF | mean | 0.474 | 0.648 | 0.563 | 0.409 | 0.650 |
| | | std.dev | 0.020 | 0.019 | 0.031 | 0.018 | 0.024 |

**Table S5.** Opportunity of equality measured by false negative rate by different models on various feature sets

| Attributed feature | HFpEF | HFrEF |
|---|---|---|
| Hispanic white | 1.230 | 0.988 |
| Sex | 1.281 | 1.030 |
| Black and White | 0.872 | 0.920 |

**Figure S1.** Workflow to identify cohort and build up ML model.

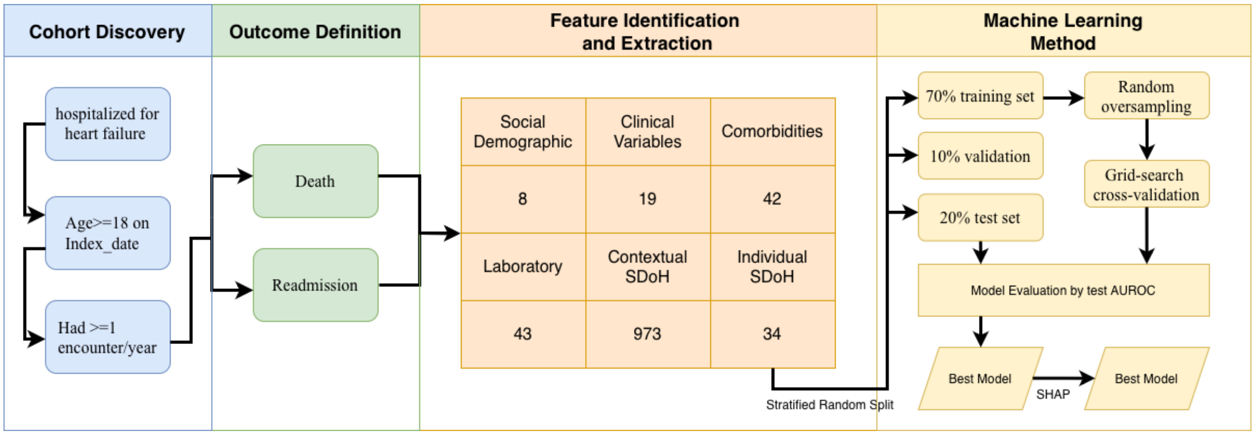


**Figure S2.** 6-month readmission/death risk by machine learning prediction risk decile using XGboost model. (a.) HFpEF population (b.) HFrEF population

**Figure S3**. SHAP values of important predictions from the original XGboost. (a.) HFpEF (b.) HFrEF.

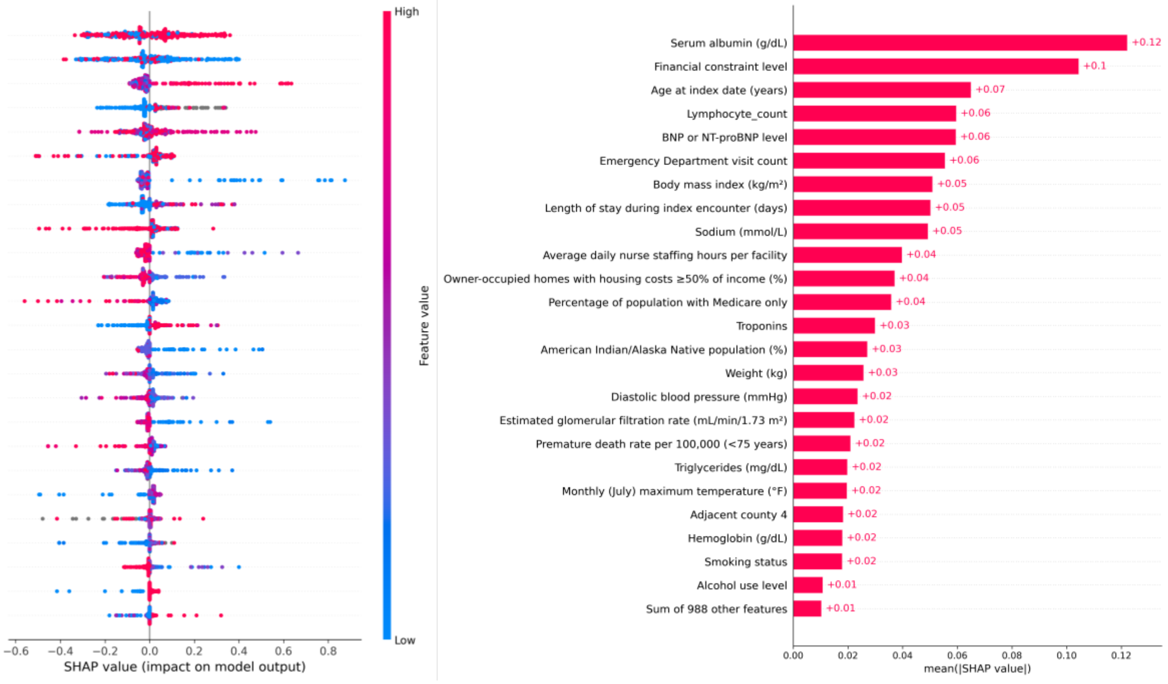


(a.)

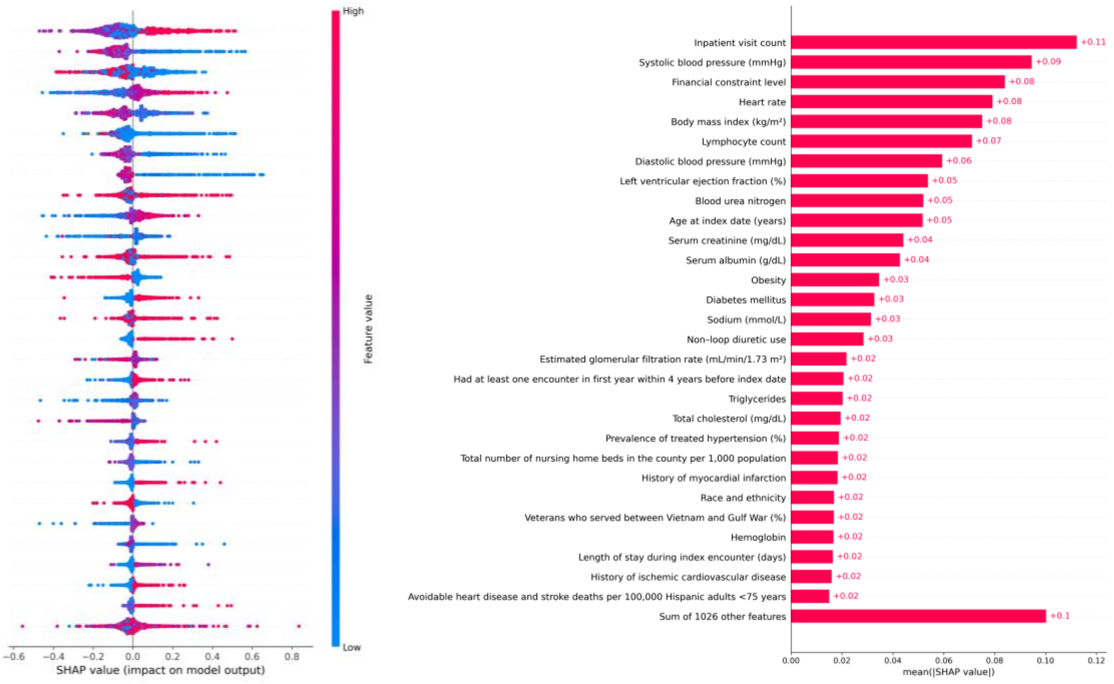


(b.)

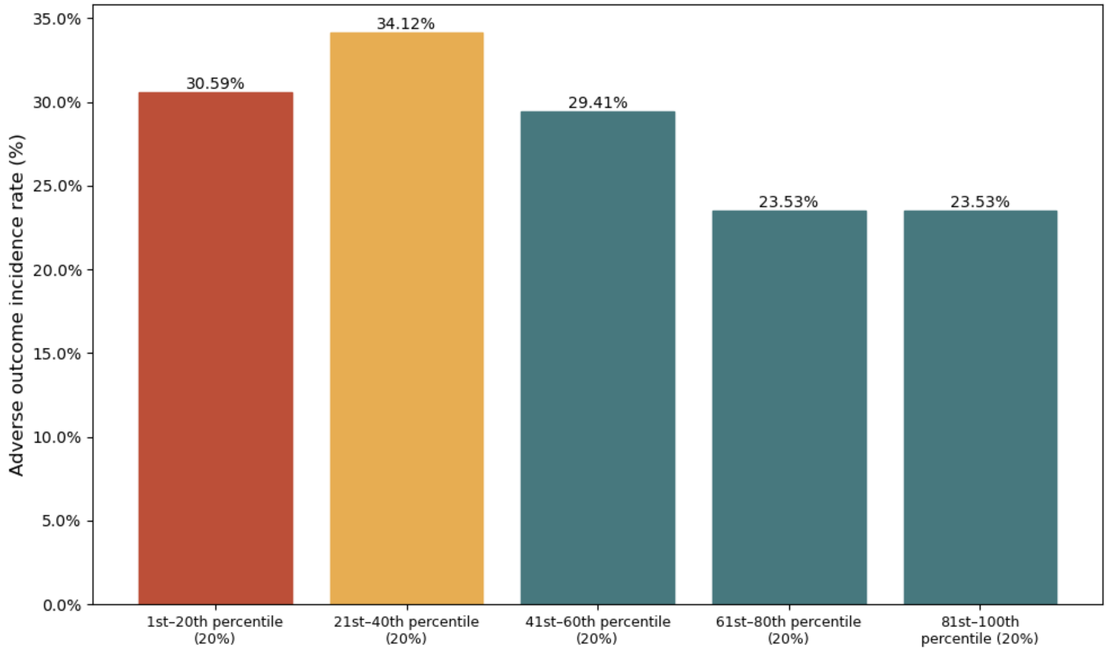


**(a)**

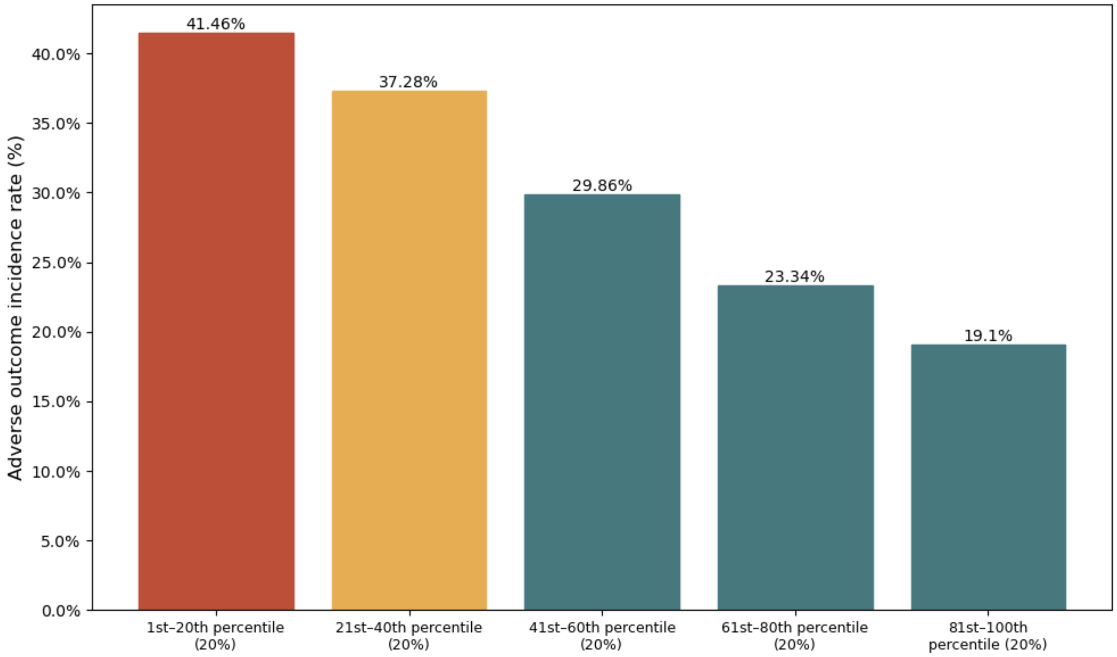


**(b.)**

**Figure S4.** The causal discovery results on full data with XGBoost model from SHAP analysis. These three images are results from CPC models. The blue nodes present SDoH and demographics variables, the green nodes stand for comorbidities and medication variables, purple nodes stand for the clinical variables, and the red node indicates the outcome. The red edges represent the indirect relationships between SDoH and outcome. (a.) HFpEF (b.) HFrEF

(a.)

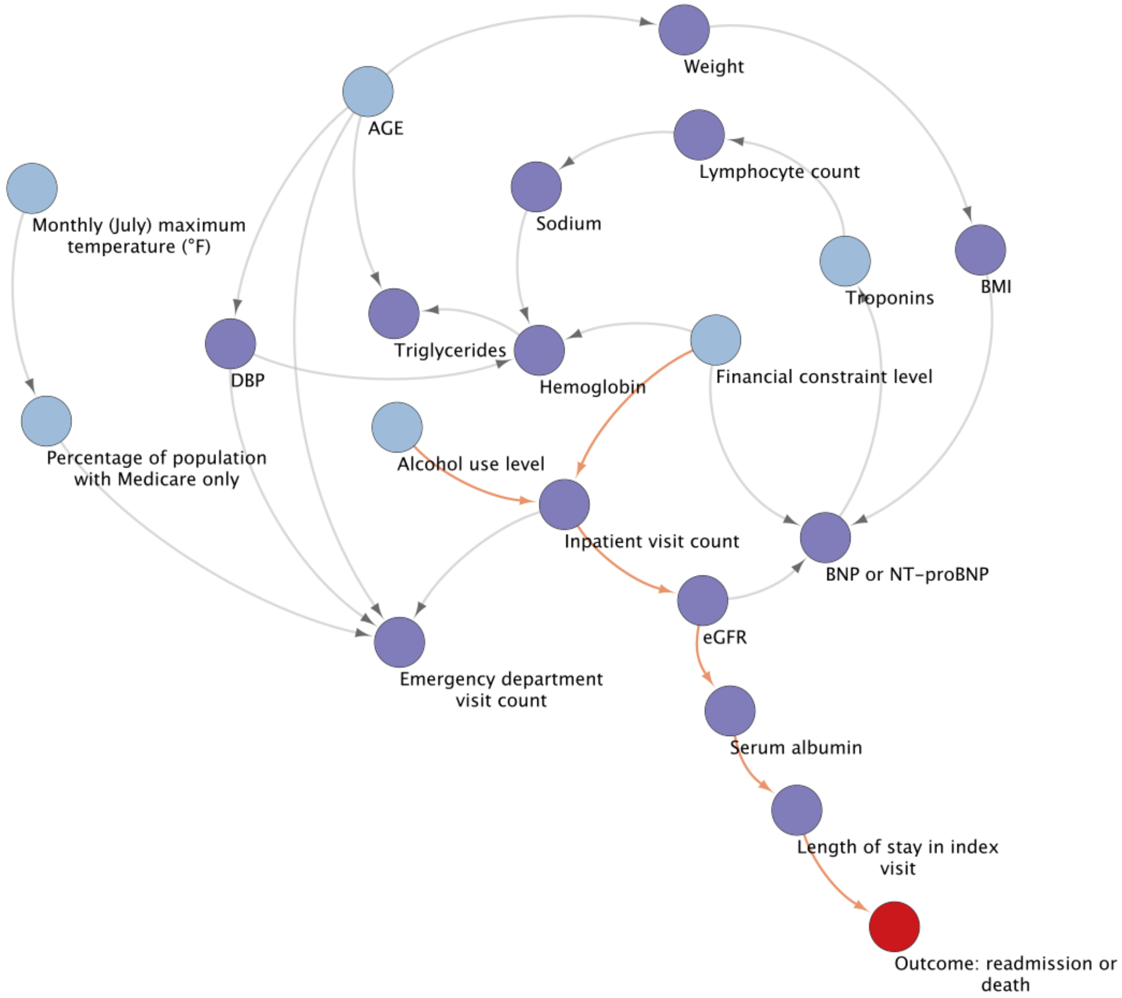

(b.)

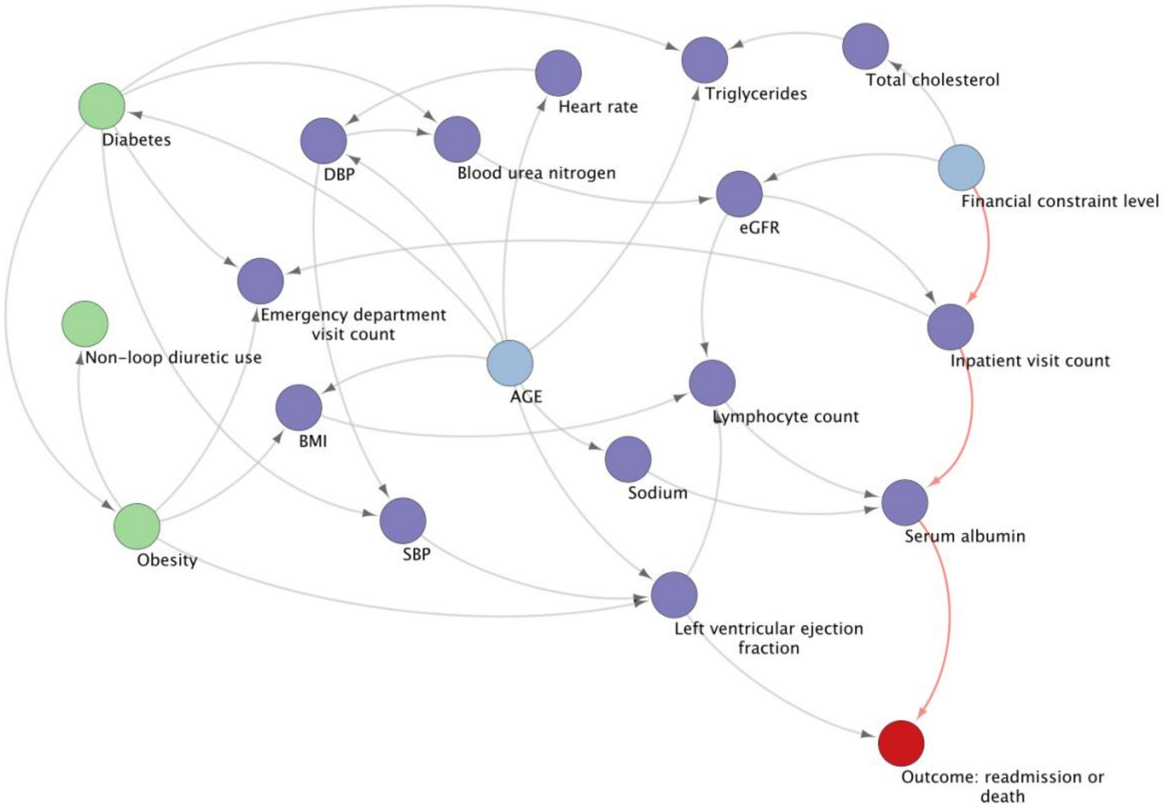


**Figure S5.** Evaluation of algorithm fairness with a focus on False negative rate disparity with XGBoost. By (a.) sex, (b.) non-Hispanic white, non-Hispanic black and Hispanic

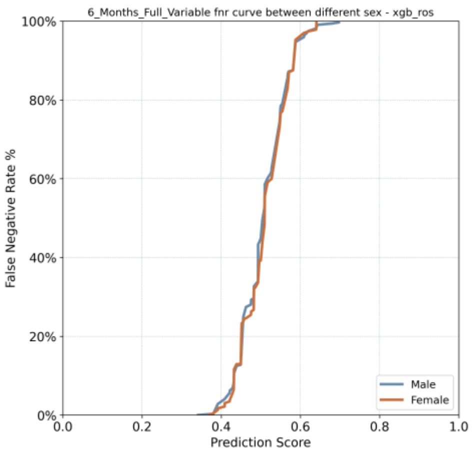


(a.)

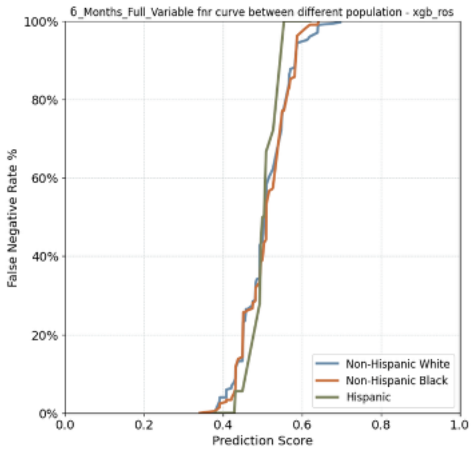


(b.)

**Figure S5 .** Impact of bias mitigation on Model Performance. The x-axis shows Equality of Opportunity, and the y-axis shows C statistic. Mitigation models reduced bias but also lowered C statistic (a.) FNR Ratio: Black/White in HFpEF (b.) FNR Ratio: Hispanic/White in HFrEF population.

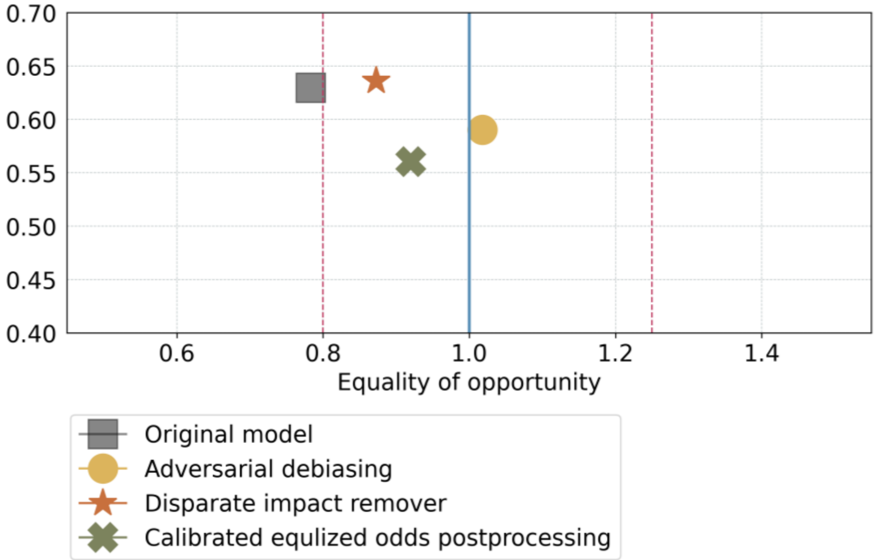


**(a.)**

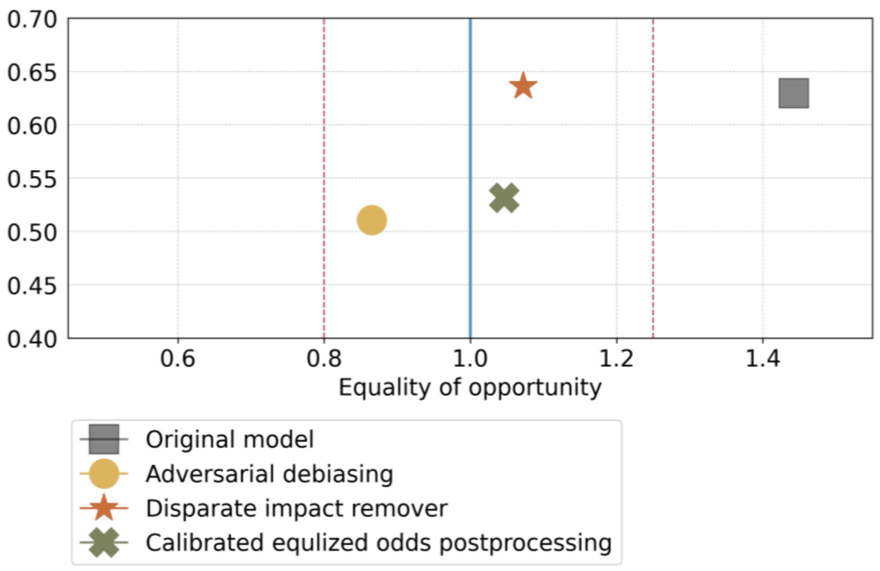
0.70
0.65
0.60
0.55
0.50
0.45
0.40
0.6
0.8
1.0
1.2
1.4
Equality of opportunity
Original model
Adversarial debiasing
Disparate impact remover
Calibrated equlized odds postprocessing

**(b.)**
